# Health Care Students’/Professionals’ Perspectives on Artificial Intelligence: Survey in Erbil, Iraq

**DOI:** 10.64898/2026.05.27.26354009

**Authors:** Aya Balisani, Dahen Zand, Naznin Virji-Babul, Tara Mohammed Ali Shallal

**Author notes:** Corresponding author: Dr. Tara Mohammed Ali Shallal., Hawler Medical University (HMU), College of Medicine, Erbil, Kurdistan Regionof Iraq.

## Abstract

**Background:** Artificial Intelligence (AI) is increasingly integrated into healthcare systems worldwide and medical schools worldwide have begun integrating AI into their curricula. The healthcare system in Iraq is currently undergoing development and AI has not yet been adopted in clinical practice in Erbil; in addition, no formal AI instruction has been incorporated into the medical education curriculum. The aim of this study was to assess knowledge levels, attitudes, and perceptions regarding AI among medical students and healthcare professionals in Erbil, Kurdistan Region of Iraq.

**Methods:** A mixed-methods survey was distributed to medical students and residents in Erbil, Kurdistan Region of Iraq. The survey was adapted from Teng et al, and modified to reflect the local context. The survey was translated into Kurdish and Arabic. Convenience sampling was used. Statistical analysis was conducted using IBM SPSS (Statistical Package for Social Sciences), Version 26.0. Chi-square and Fisher’s exact tests were used to test associations between categorical variables. Mann Whitney U test was used to compare mean ranks between groups in the non-normally distributed data. A P value <0.05 was considered statistically significant. Thematic analysis was applied to open-ended qualitative responses by two independent reviewers.

**Results:** A total of 368 participants participated in this study. The majority (85.6%) of participants felt that AI should be taught in schools and universities, and 90.8% reported using AI. ChatGPT was by far the most commonly used AI tool (85.3%). Participants aged 20-24 years (93.2%) and 25-29 years (90.2%) showed the highest prevalence of using AI. Participants that used AI previously, had higher scores for support for AI development in their field (U = 3744.5, P=0.001), feelings of hope towards AI in their field (U = 4406.5, P = 0.004) and thinking that students should learn the basics of AI (U = 4022.5, P = 0.03). Male participants were more likely to use AI in comparision with women (P=0.045). The most common concern regarding AI was loss of jobs (33.0%), followed by overreliance on AI (22.8%). Qualitative analysis revealed themes of guarded optimism, and concerns regarding the ethical implications of AI use in medicine.

**Conclusion:** Medical students and physicians in Erbil are early adopters of AI in spite of any formal training. In parralel, most participants expressed dissatisfaction with their understanding of the ethical implications of AI in healthcare and emphasized the need for formal AI education in healthcare curricula. The majority of participants expressed guarded optimism regarding the future of AI in healthcare. A gender gap in AI was identified, consistent with global trends with implications for professional equity.

## Introduction

### Background

Artificial Intelligence (AI) is increasingly being integrated into healthcare systems worldwide, influencing clinical decision-making, patient care and diagnostics.^1^ AI technologies have been applied in a wide range of medical domains, including providing the highest standard of patient care, streamline clinical workflow and improve diagnostics.^2^ Several medical specialties including radiology, pathology, and dermatology have already demonstrated substantial potential for AI integration.^3,4^ As AI becomes more widely incorporated into healthcare systems, healthcare professioinals are increasingly expected to understand their capabilities, limitations, and ethical implications in order to use them safely and effectively in clinical practice.

In many countries, AI-based technologies are already being integrated into clinical workflows. In countries such as France, Hungary, Portugal, the Netherlands, and Sweden, these tools are used to support diagnostic decision-making, patient support through chatbot technologies, and the use of AI to enhance the efficiency of administrative and logistical processes.^5^. In parrallel, healthcare systems facing workforce shortages and increasing patient demands have explored AI applications to improve efficiency, expand virtual care, and support clinical services.^6^

#### AI in Medical Education

Beyond clinical practice, AI is also reshaping how future healthcare professionals are trained. Medical schools worldwide have begun integrating AI into their curricula, recognizing the need to prepare future physicians to work alongside these technologies. Many medical schools have been incorporating AI related content with the aim of equipping students with foundational knowledge in machine learning, improving understanding of how AI algorithms function, and the skills required to critically evaluate AI-generated outputs in clinical practice. ^12,13^

The potential applications of AI in medical education are extensive and address several longstanding challenges in traditional medical training. These challenges include limited student engagement associated with lecture-based instruction, the increasing volume and complexity of medical knowledge, and variability in specialty training that may result in educational inconsistencies.¹⁴ Key applications of AI in medical education include virtual simulations and augmented reality technologies, which are increasingly used to create immersive and realistic clinical, anatomical, and surgical scenarios, enabling students to practice and refine their skills in a safe environment without risk to patients. In parallel, adaptive learning platforms deliver personalized educational experiences by dynamically adjusting the difficulty and content of materials in response to individual student performance and learning needs. Natural language processing tools, including chatbots, further support student learning by answering questions and aiding in the understanding of complex medical concepts. Additionally, AI-based assessment and evaluation systems assist educators by supporting the grading of assignments, assessing clinical competencies, and identifying students who may be at risk of academic underperformance or attrition.^14^

#### Healthcare system in Erbil/Iraq

The healthcare system in Iraq continues to undergo development, although digitization and AI integration remain limited. In Erbil, a city in northern Iraq, there are 59 hospitals, comprising both public and private institutions.^7^ Hospital capacities in Erbil vary, ranging from smaller hospitals with 40-bed capacities to larger institutions accommodating up to 10,000 patients.^8^ Overall, Erbil has a capacity of approximately 2 hospital beds per 1,000 population.^9^ The majority of hospital admissions in Erbil are related to non-communicable diseases,^10^ with other common causes including trauma and communicable diseases.

Despite the growing interest in technological advancements, most hospitals and clinics in Erbil have yet to digitize patient records. ^11^ Currently, patient histories, operation notes, medication charts, follow-up documentation, and discharge summaries are all handwritten by either doctors or nurses.

At present, AI is not utilized in clinical practice in Erbil, despite its growing adoption in healthcare systems globally. This lag in implementation, however, does not imply that healthcare professionals and students in Erbil are unaware of or uninterested in these technologies. While AI has not been formally integrated into medical training, educational methods in the region have largely embraced digitalization. Professors typically provide lectures in digital format such as PowerPoint presentations and PDFs, which students access through tablets, smartphones, and laptops. This contrast in the use technology between university and hospital settings may be attributable to several factors. Although data on this issue is limited, it is plausible that transitioning to electronic lecture formats requires less training and adaptation than implementing electronic databases for patient management. Furthermore, universities often offer workshops and training sessions to help staff adjust to technological advancements, whereas such support is generally lacking in hospitals.

In recent years, there has been a notable increase in students’ interest in AI applications, with many turning to tools like ChatGPT to assist with writing, problem-solving, and completing assignments. While there is limited data on this phenomenon in Erbil specifically, a study conducted at a university in Baghdad revealed a generally positive attitude among students toward incorporating ChatGPT into their educational practices.^15^

This informal adoption of AI tools among students in Erbil, occurring without formal guidance or ethical frameworks, mirrors patterns observed in other settings where technology access has outpaced institutional adaptation. However, the specific context of Erbil, where students will transition from digitally-enhanced education to largely paper-based clinical practice, creates unique challenges for workforce preparation. Despite the growing interest, no formal AI classes have been integrated into the medical education curriculum, nor have AI tools been utilized for student learning formally.

### Rationale and Aim

Although AI has not yet been widely adopted within Iraq’s healthcare system or medical education, rapid global advancements suggest that its integration is inevitable. To ensure future readiness, it is essential that medical students and healthcare professionals develop an informed understanding of AI applications, limitations, and ethical implications. Establishing a baseline assessment of knowledge, attitudes, and perceptions toward AI is a critical step in guiding curriculum development and targeted educational interventions.

Despite the growing interest and informal use of AI tools among students, no formal AI classes have been integrated into the medical education curriculum, nor have AI tools been utilized for student learning in a structured manner. This disconnect between informal adoption and formal training creates an urgent need to understand current knowledge levels and attitudes.

Therefore, the aim of this study was to assess the level of knowledge, as well as the attitudes and perceptions regarding AI among medical students and healthcare professionals in Erbil, Kurdistan Region of Iraq.

## Methods

### Study Design and Setting

This study was a cross-sectional study conducted in Erbil, Kurdistan Region of Iraq.

#### Data Collection

We adapted a survey previously used by Teng et al.¹^6^, modifying some questions to better fit the Iraqi context. The survey was translated into Kurdish by co-author D. Zand and into Arabic by first author, A. Balisani. It was distributed in online Google Forms for doctors and students. The survey consisted of three sections: The first addressing demographic information (age, gender, stage of career), the second regarding AI use (agent used, assessing experience/comfort with forming AI prompts), and the third regarded perceptions and attitudes towards AI, including Likert-scale questions. A pilot was first carried out on 20 individuals to determine potential areas of improvement. The data was collected over a 4 month period from July 2024 to October 2024.

#### Sample Size and Selection Criteria

Participants were recruited using convenience sampling. Our goal was to obtain a large, diverse, and inclusive sample that accurately represents the broader population. The inclusion criteria were medical students and medical graduates. Exclusion criteria were medical school graduates who did not do clinical work and pursued other careers. Participants were recruited through several channels: class representatives and teachers distributed the survey to students, colleagues were asked to share the survey link within their professional networks, and the link was also be posted in shared social media group chats for specific professions. Additionally, physical copies of the survey were distributed to healthcare workers in a selection of randomly chosen hospitals and clinics in Erbil, such as Balsam Private Hospital, Mihrabani Private Hospital, Zanko Private Hospital, and Dr. Shwan’s Cardiology Clinic.

#### Ethics

Ethical approval was obtained from the Bioethics Committee at the College of Medicine, Hawler Medical University (HMU). In order to ensure confidentiality, a master list of code numbers were employed to replace the subject’s name on all documents and labels and in all experimenter discussions to protect the identity of each participant. A unique alphanumeric number, not derived from personal identifiers was used. The only identifying information that the researcher collected was the consent form.

The survey was conducted using either an anonymous online platform or anonymized physical copies. Collected data was stored in Google Docs for approximately four months, until data analysis began.

#### Data Analysis

Data was analyzed using IBM SPSS Statistics, Version 26.0. Categorical data was demonstrated as frequencies and percentages. Normality testing (the Shapiro–Wilk test) was conducted for the numerical data, normally distributed data was demonstrated as means ± standard deviations while non-normally distributed data was demonstrated as medians (interquartile ranges (IQR)). Normality test for the attitue score variables could not be assessed accurately (W=0.913, P <0.001). Therefore, Mann whitney U tests were conducted to assess difference of the scores between groups that used AI and those that did not. Chi-square tests were applied to categorical data. Statistical significance was considered at p-values < 0.05. Thematic analysis was employed in the qualitative section to determine the general themes found in participants’ responses. In addition, to ensure rigor and consistency of the qualitative analysis, AB and DZ independently reviewed all 206 open ended responses. Each generated initial codes and proposed themes independently. They then met to compare their analyses, discuss any discrepancies, and reach consensus on the final themes.

The data was used only for this research project and was not disseminated publicly, except in summary form through professional journals or books or scholarly meetings. As per the ethics protocol, individual subjects were not identified and only aggregated data were used for presentation and publication.

## Results

### Participant demographics

Our study included 368 participants, with a mean age of 27.2 ± 8.0 years. The demographic details are summarized in Table 1 and Figs. 1 and 2.

**Figure 1:**
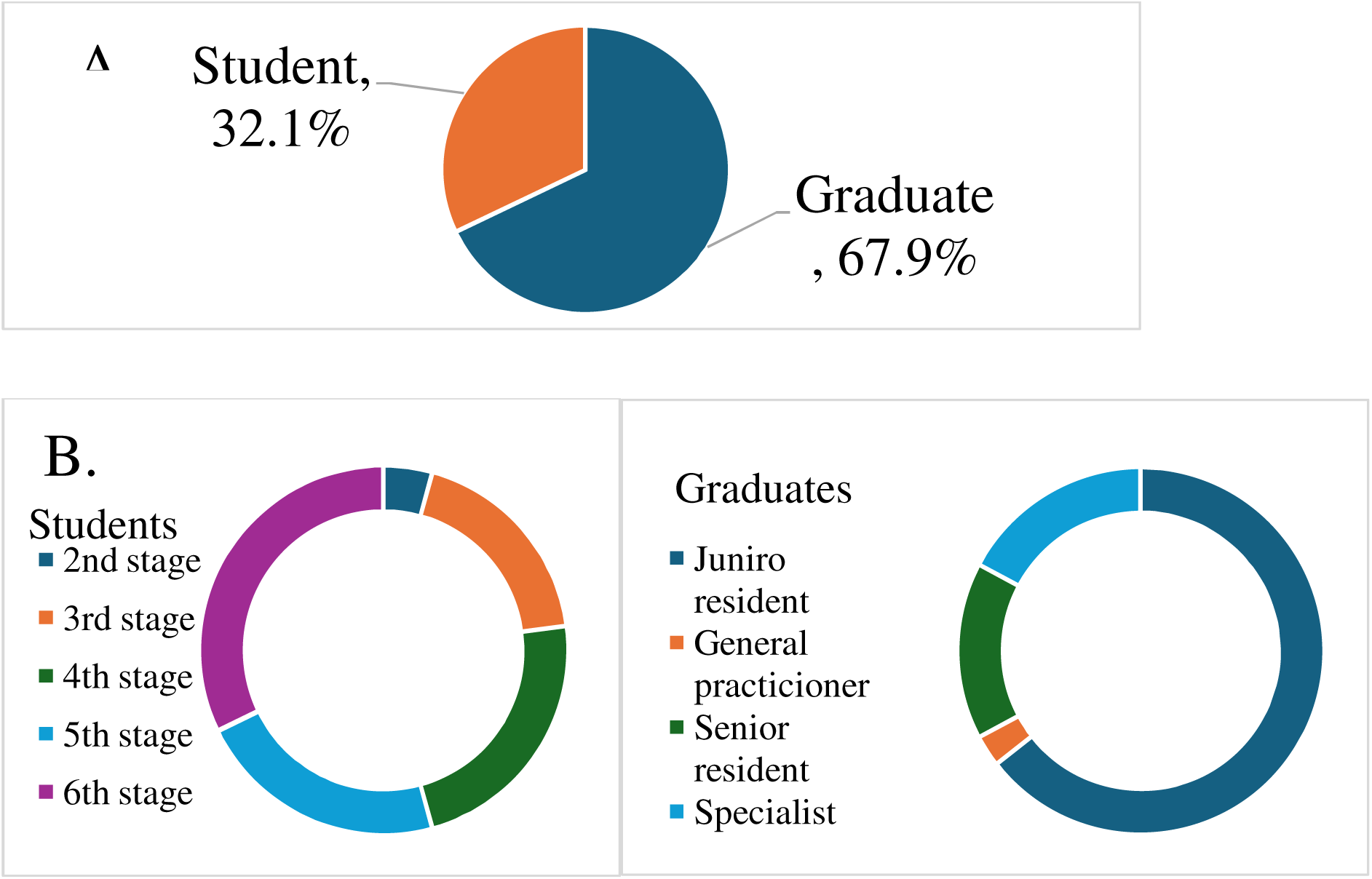
Stage of Career of Study Participants. 1A Breakdown of whether the Participants is a Student or Graduate. 1B. Breakdown of Stage of Students and Graduates.

**Figure 2.**
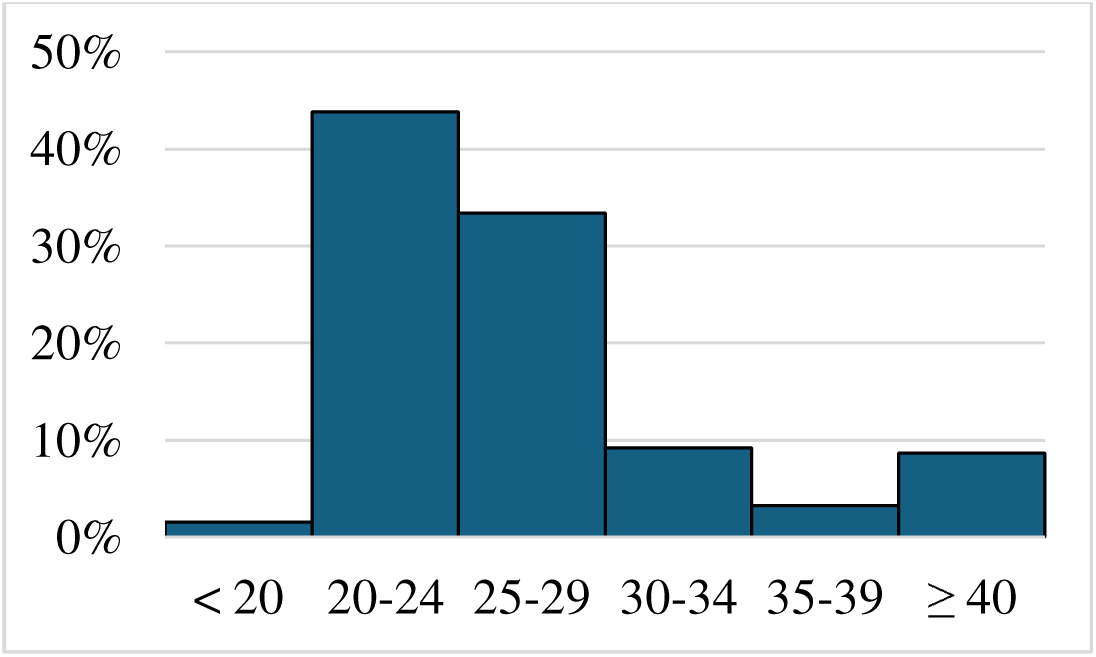
Age Distribution of Study Participants.

**Table 1.** Demographics of the Study Participants.

| Characteristic | Frequency (n.) | Percentage (%) |
| --- | --- | --- |
| Gender |  |  |
| Female | 178 | 48.4 |
| Male | 190 | 51.6 |
| Highest degree of education |  |  |
| Bachelor's degree | 210 | 57.1 |
| Board certification | 29 | 7.9 |
| Diploma | 1 | 0.3 |
| High school | 107 | 29.1 |
| Master's degree | 10 | 2.7 |
| PhD degree | 11 | 3.0 |

Career aspirations of the participants were evaluated, and the results are summarized in Figure 3.

**Figure 3.**
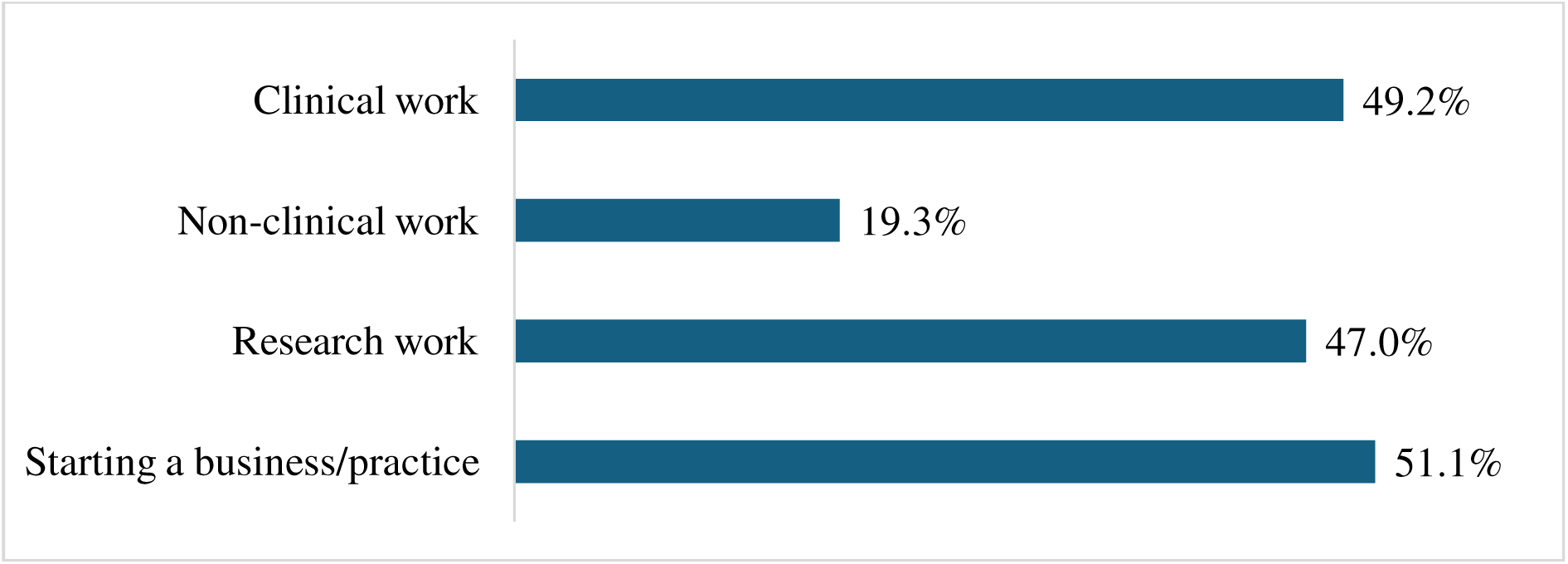
Career Aspirations of Study Participants

### Attitudes about AI

Overall, 85.6% of the participants indicated that AI should be taught in schools and universities. Participants were asked to rate from a scale of 1 to 10 how much they agreed with various statements regarding AI. Their responses are summarized in Table 2.

**Table 2.**
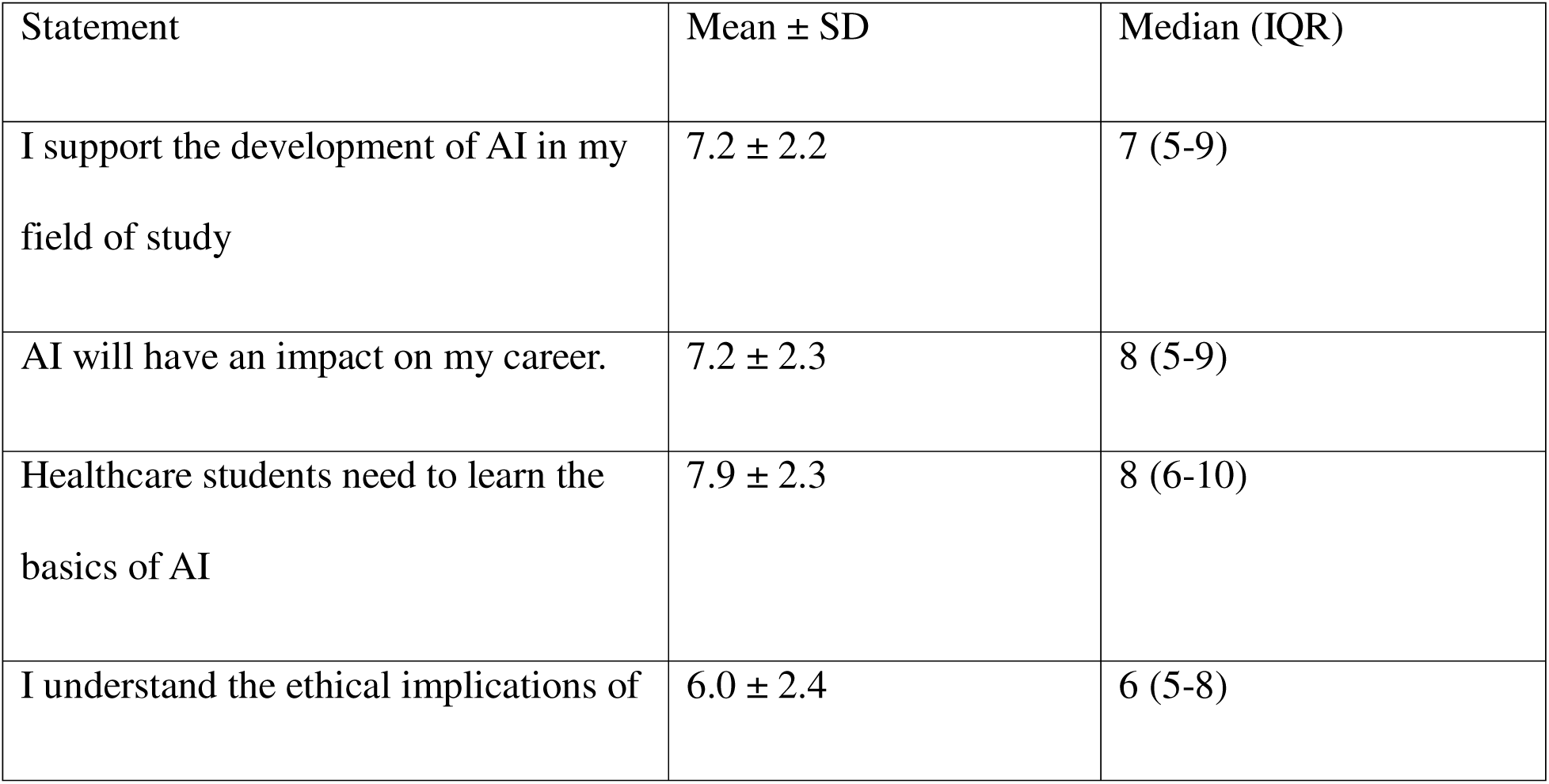
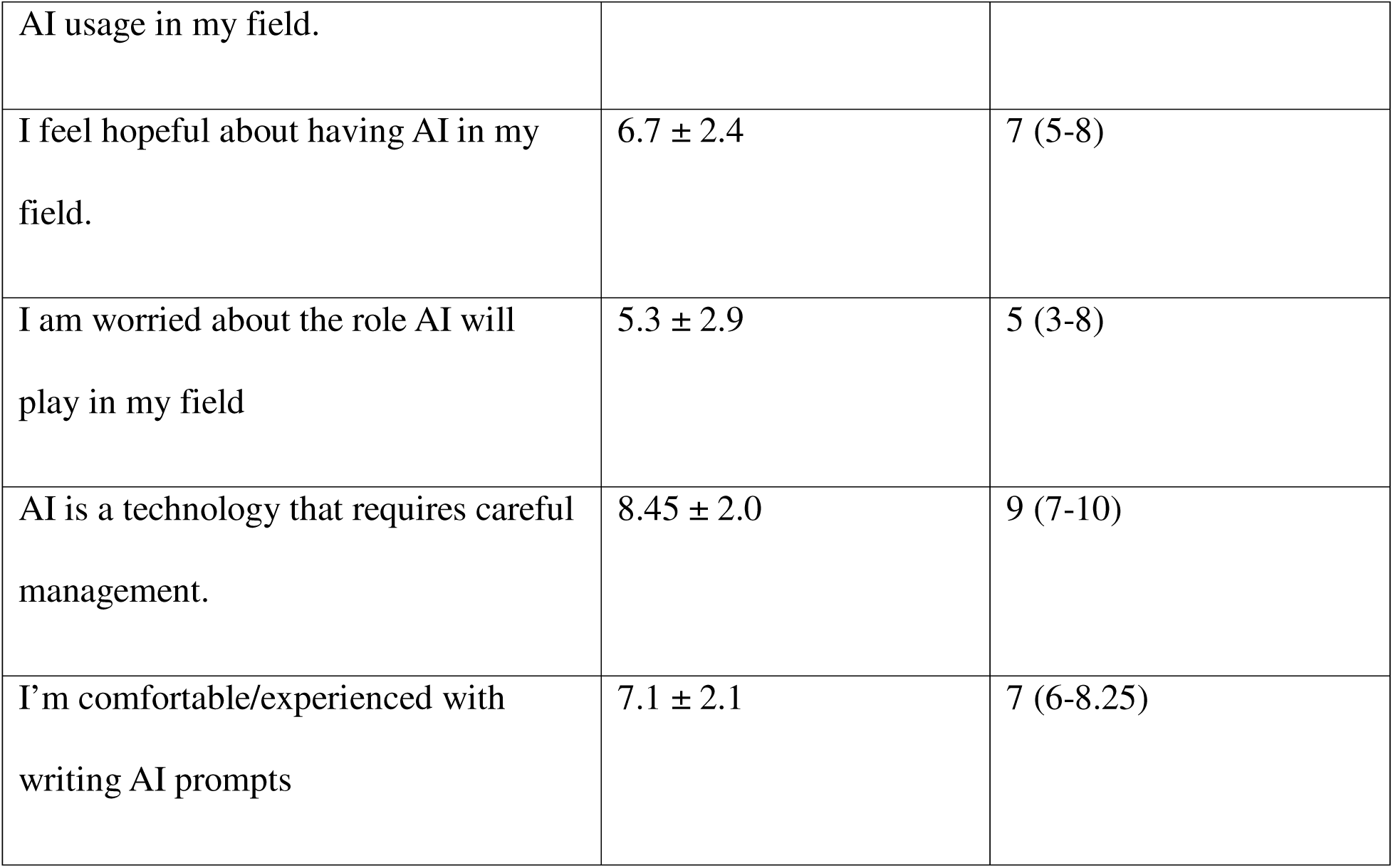
Mean ± Standard Deviation (SD) and Median (Interquartile Range) of Participant Responses on a Scale of 1 to 10 to Statements Regarding AI.

Furthermore, the majority of participants (90.8%) reported using AI. The distribution of AI use by age is shown in Figure 4. The types of AI used by the participants is shown in Figure 5.

**Figure 4.**
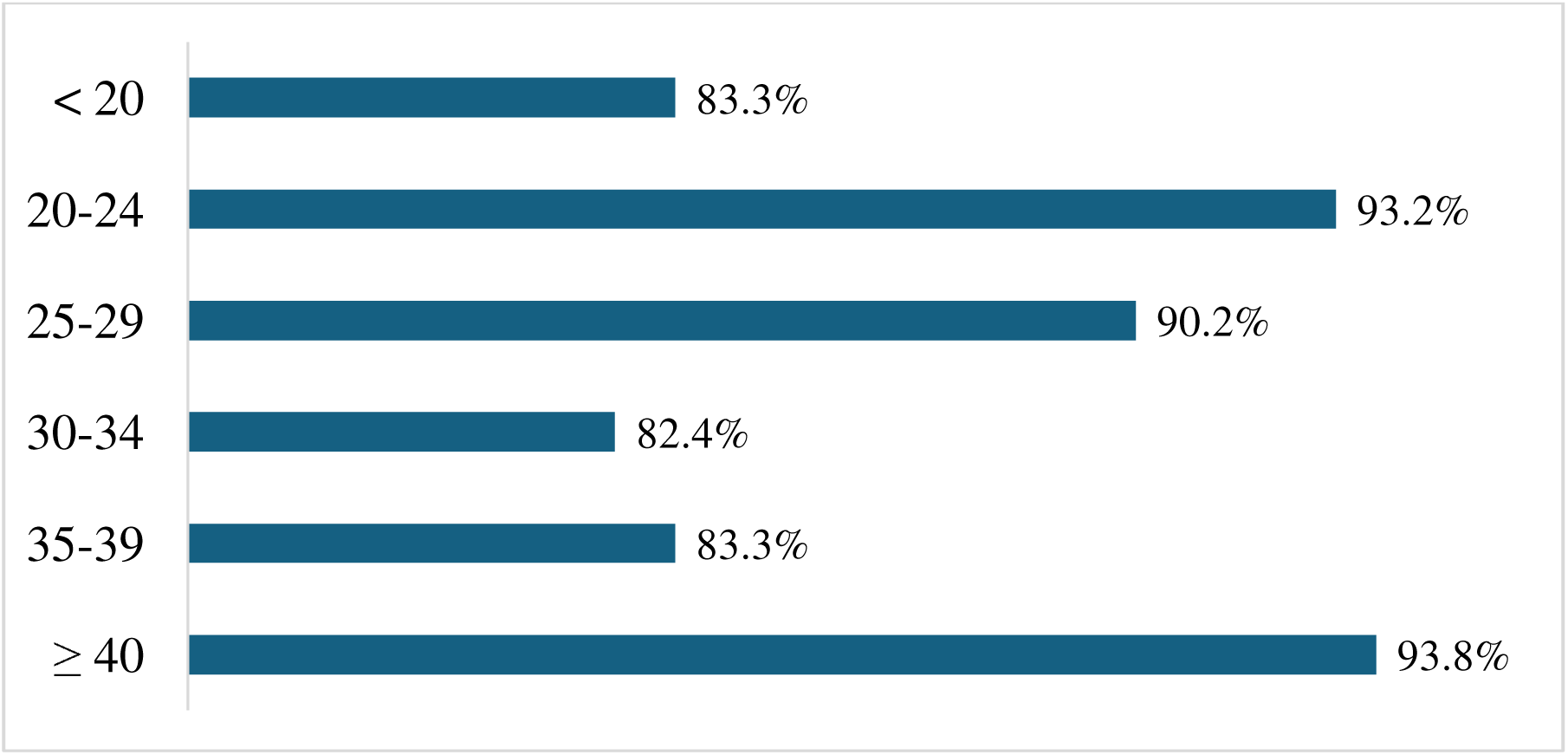
Prevalence of AI Use in Different Age Groups.

**Figure 5.**
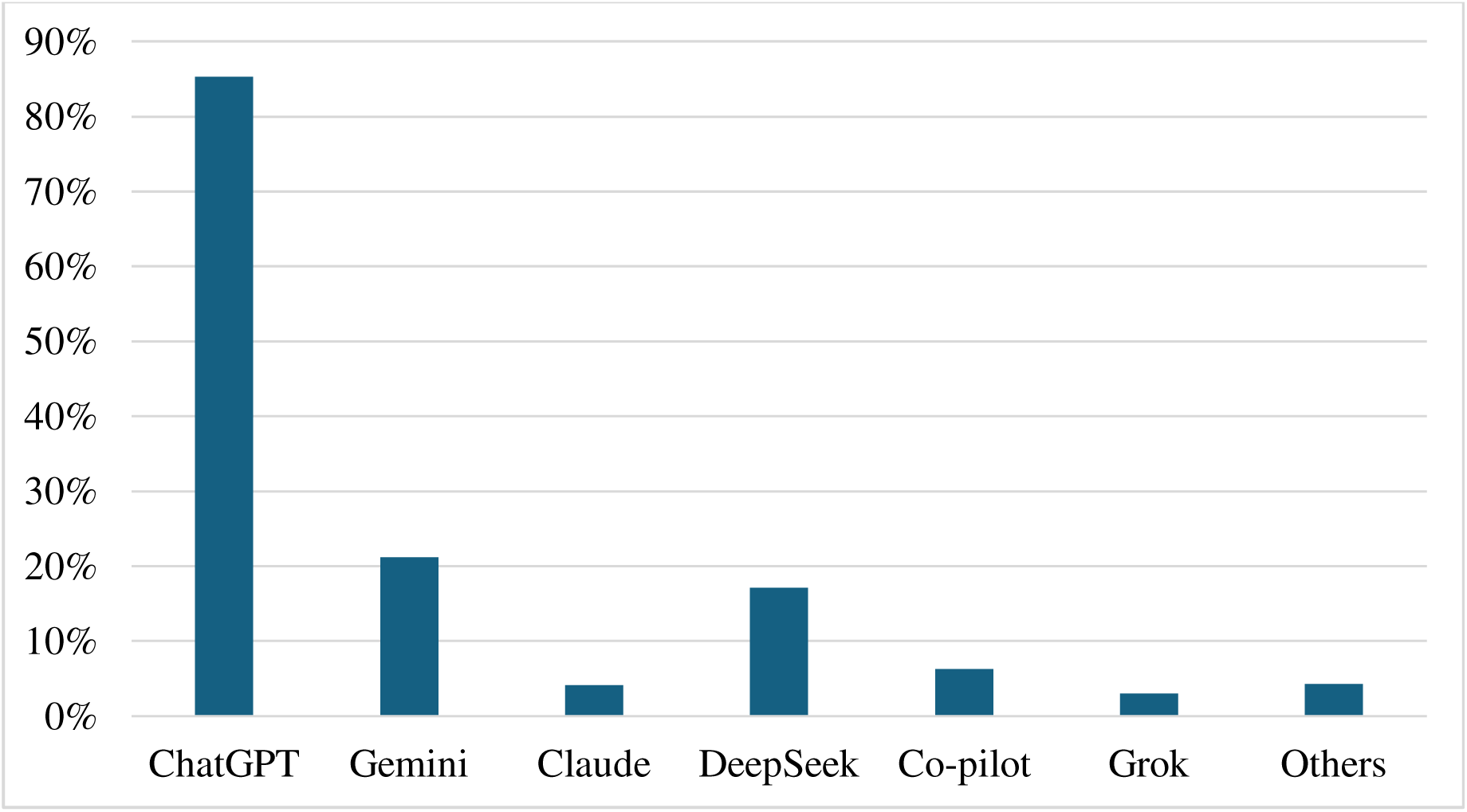
Prevalence of Use of Different AI Models among Study Participants.

Mann Whiteny U tests were conducted to compare the attitude scores between individuals who used AI and those that did not. The results revealed a significant difference between the medians of support for AI (U = 3744.5, P=0.001), thinking that healthcare students need to learn the basics of AI (U = 4022.5, P = 0.004) and feeling hopeful about AI (U = 4406.5, P = 0.03) between the two groups. There were no statistically significant differences between the rest of the attitude scale responses and the use of AI. These findings are illustrated in Figure 6.

**Figure 6.**
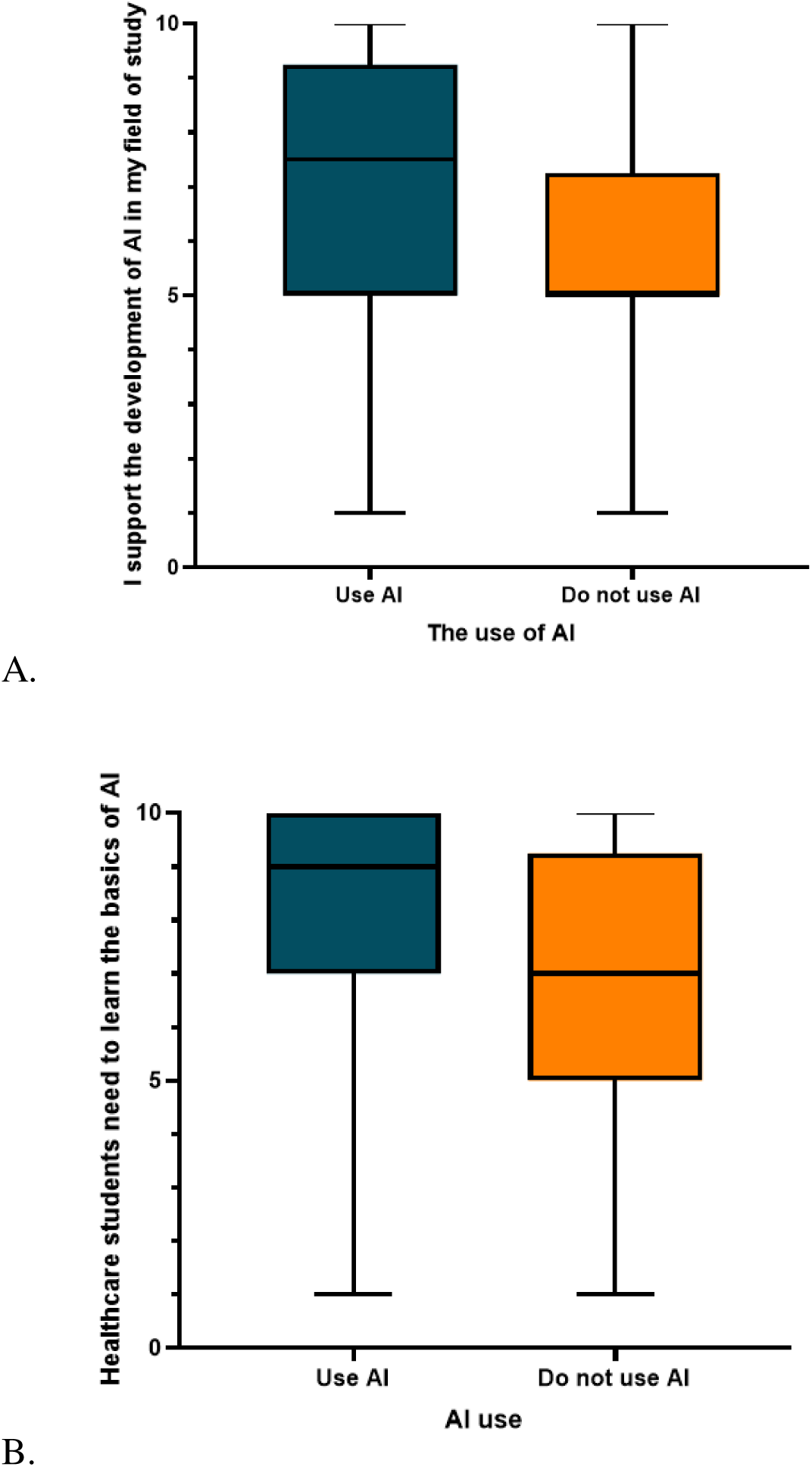
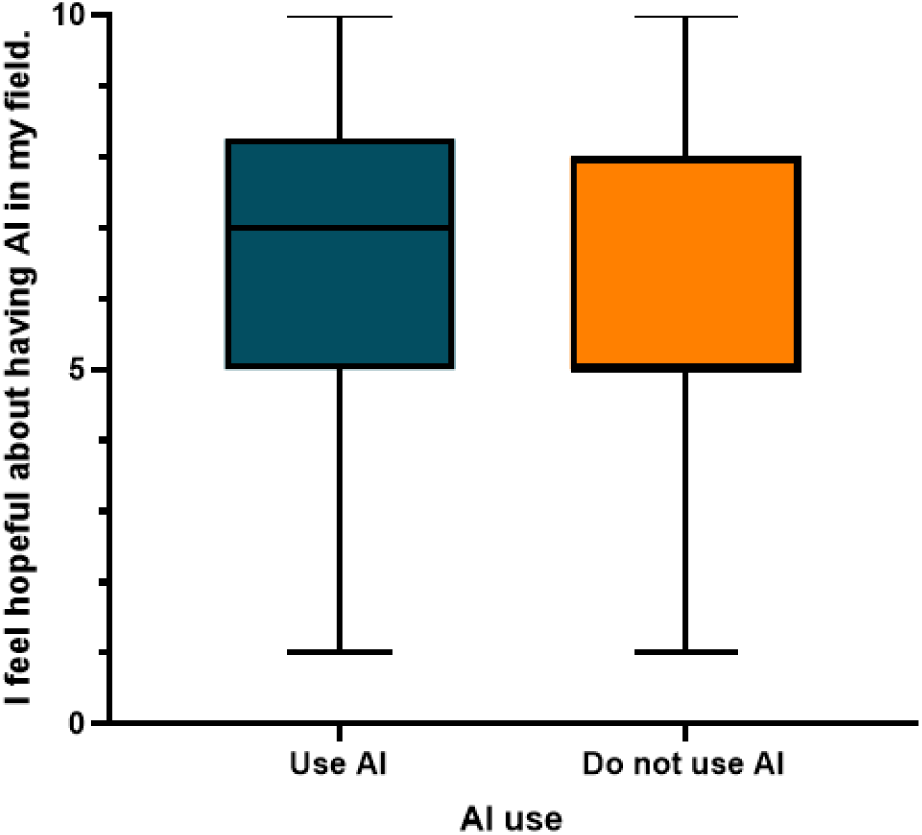
Comparing the Attitude Scores between those that Use AI and those that did not.

Chi-square analyses were performed to determine the associations between various subject characteristics and AI use as well as thinking that AI should be taught in schools universities. We found gender, wanting to pursue one’s own business/practice to be significantly associated with AI use. Furthermore, wanting to pursue clinical work and one’s own business/practice both had associations with one’s opinions regarding the teaching of AI in schools and universities. These are summarized in Tables 3 and 4.

**Table 3.** Association between Gender and Wanting to Start a Practice/Business with the Use of AI.

| Characteristic | AI use |  | P-value |
| --- | --- | --- | --- |
|  | No, n. (%) | Yes, n. (%) |  |
| Gender |  |  |  |
| Female | 22 (12.4) | 156 (87.6) | 0.045 |
| Male | 12 (6.3) | 178 (93.7) |  |
| I want to start my own practice/business |  |  |  |
| No | 11 (6.1) | 169 (93.9) | 0.043 |
| Yes | 23 (12.2) | 165 (87.8) |  |

**Table 4.** Association between Wanting to Pursue Clinical Work and Wanting to Start a Practice/Business with Believing AI should be Taught in Schools and Universities.

| Characteristic | AI should be taught in schools/universities |  | P-value |
| --- | --- | --- | --- |
|  | No, n. (%) | Yes, n. (%) |  |
| I want to do clinical work |  |  |  |
| No | 19 (10.2) | 168 (89.8) | 0.018 |
| Yes | 34 (18.8) | 147 (81.2) |  |
| I want to start my own practice/business |  |  |  |
| No | 36 (20.0) | 144 (80.0) | 0.003 |
| Yes | 17 (9.0) | 171 (91.0) |  |

Participants were asked about the ethical considerations regarding AI and to respond with their concerns for AI through open-ended questions. 206 participants provided usable answers to this prompt. The most prevalent concern was the loss of job opportunities for medical professionals, followed by medical professionals becoming over-reliant on AI. These are demonstrated in Figure 7.

**Figure 7.**
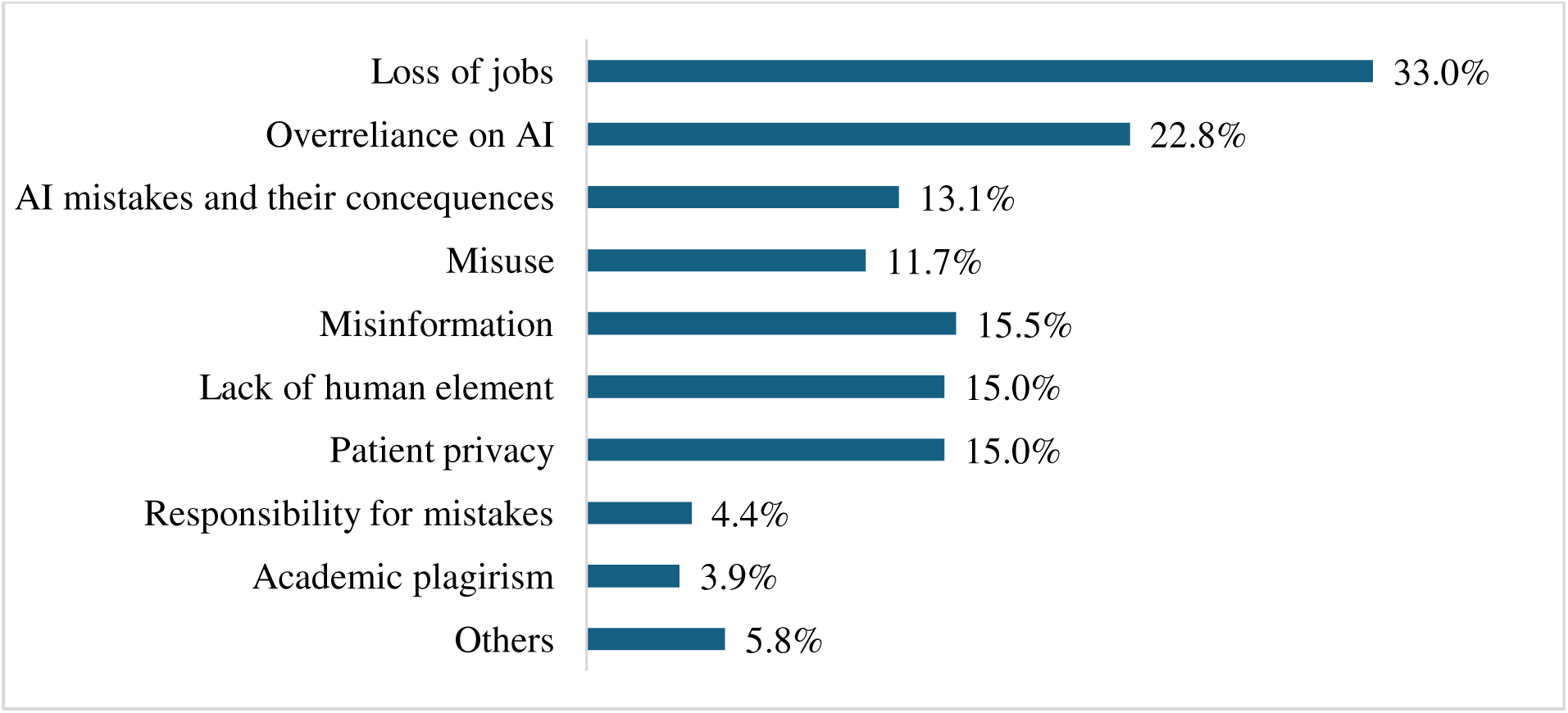
Concerns and Ethical Considerations Regarding AI among Study Participants

### Qualitative data analysis

To contextualize the quantitative findings and explore the reasoning behind participants’ attitudes, we analyzed 206 open-ended responses (56% of the total sample) regarding their hopes, concerns, and ethical considerations about AI in medicine. The qualitative analysis revealed 2 key themes: Guarded optimism and concerns and ethical implications.

#### 1. Guarded Optimism

More than half of the study participants, spanning all ages and career stages, expressed a sense of hope regarding the role AI may play in their field. Reflections such as “*Helps me learn at a significantly faster rate, make fewer mistakes, do most of the repetitive, boring tasks for me so I can focus on the more important aspects of medicine,*” reported by a 50-60-year-old specialist, and “*It can help me stay updated about recent guidelines and having fast access to knowledge*.” reported by a junior house officer in their early 20’s, illustrate a broadly optimistic outlook toward AI across generations.

Medical students, in particular, conveyed considerable enthusiasm, both for AI’s potential to enhance their education and to support their future careers. For instance, one 3rd year student stated, “*It would definitely increase the speed of development of research, diagnostics, treatment plans and their efficiency.”* A 6th year student noted, “*AI algorithms excel at analyzing medical images like ultrasounds, MRIs, and mammograms. They can detect subtle patterns that might be missed by the human eye*…” Similarly, a 4th year student shared, “*There are many topics in medicine that are hard to understand but AI makes them easier*,” while a 5th year student emphasized, “*AI can make the task of gathering information from different sources and medical books easier*.”

Despite this optimism, some students expressed reservations, most notably regarding the possibility of AI replacing physicians. One 5th year student remarked, “*I’m not sure in one hand it will be helpful but also I’m worried it will replace my role as physician.*” A 6th year student stated, “*Not hopeful, I don’t find it useful*!” while another 6th year student commented, “*Yes, it helps us in many ways, but at the same time the thought of replacing us makes me worry*.”

Recently graduated doctors working as junior house officers (JHOs) and senior house officers (SHOs) similarly demonstrated optimism, particularly regarding AI’s potential to reduce workload, save time, and streamline clinical responsibilities. Illustrative responses include: “*It aids me in doing difficult tasks easily*.” (JHO); “*It would aid medical care and especially diagnosis and reduce the load on healthcare workers*.” (JHO); “*Due to large amount of information, I needed to memorize regarding different medicines, it could be confusing sometimes, so having AI with being a reminder is just hopeful to prevent side effects or negative consequences.” (SHO); and “It helps you to not waste your time on useless things*.” (SHO).

Nevertheless, caution was also evident among medical graduates. Concerns centered on the reliability of AI-generated information and its potential impact on professional development. One JHO stated, “*I never trust anything other than human beings to be Healthcare provider*,” while an SHO reflected, “*It may help me now but it can make me dumber since I don’t do as much research as before so I can only assume how it is for the others*.”

Specialists likewise expressed optimism about AI’s integration into clinical practice. An orthopedic surgeon noted, “*Because it gives the prospect of better healthcare to people.*” A pediatrician commented, “*It will help with the quick and plain decisions that need to be taken*,” while a rheumatologist observed, “*Makes tasks easier and more manageable*.” An anesthesiologist added, “*Yeah it really has helped me practically in revising some critical information and summarizing important topics for me. It’s literally like a friend to me*.”

At the same time, specialists articulated measured concerns. A pediatric surgeon stated, “*Can’t tell a definite answer, it depends on putting regulations for using it*,” reflecting the importance of governance and oversight. A nephrologist described their stance as “*neutral, I’m not sure about all the benefits it can have*,” while a general surgeon cautioned, “*May be harmful for people if they use it without full information and awareness*.”

Overall, the findings reveal a prevailing sense of optimism toward AI across all professional stages, tempered by thoughtful concerns regarding regulation, reliability, and the preservation of the physician’s role.

However, caution and worry was also expressed by half of the participants, and we can see that through statements such as “our roles as doctors are threatened”, a 5^th^ year medical student and “*I’m worried it’ll lead to a loss of job opportunities*”, a junior house officer and “*It may take our jobs as doctors*” a pediatric surgeon. The common theme of concern was loss of jobs.

#### 2. Concerns and Ethical Implications

Many participants expressed significant concern about the ethical challenges posed by the use of AI in medicine. Issues related to patient privacy and the risk of misinformation emerged as particularly prominent. As one third-year medical student noted, “*I cannot rely on only AI’s information and advice for the health of people,*” while a fourth-year student emphasized that “*Privacy of data will be in question*.” These concerns were echoed by a fifth-year medical student, who stated that “*using AI in medicine affects patient privacy, safety, and trust. Doctors must make sure AI tools are accurate, fair, and used in the right way, without replacing human judgment*.”

Junior doctors voiced similar apprehensions, particularly regarding the potential for AI-generated misinformation to cause patient harm and the risk of breaches in confidentiality. As one explained, “*Because it involves patient privacy, informed consent, data security, and ensuring AI supports not replaces human judgment*.” Another junior doctor highlighted the professional implications of overreliance on AI, stating, “*Depending on AI and only believing in the information it provides means not believing in your own knowledge as a doctor, and sometimes this knowledge determines the fate of someone*.” Such perspectives were commonly shared among junior doctors.

Additionally, many junior doctors raised concerns about accountability in the event of AI-related errors. One JHO questioned, “*For example what if the AI makes a mistake, then who would be responsible?”*

Among SHOs and specialists, comparable concerns were expressed regarding data privacy and patient safety. One participant cautioned that “*Entering the data and information of the patient into AI can compromise their privacy,*” while another warned that “*Private data is not secure on AI servers. Also, Using AI guidance without fact checking leads to errors and malpractice*.”

In contrast, a minority of participants perceived fewer ethical concerns associated with AI use. One junior doctor reflected, “*It might be seen that ethically that doctors should not use AI they should depend on their information to treat the patient, but I don’t see it like that, few decades ago doctors were only using their hands and few devices to diagnose the patients, nowadays they use a lot of investigations, then what is wrong with development and using AI to serve the benefit of patients*?” Similarly, a pediatric surgeon described a supportive role for AI, stating, “*I use it for self-reassurance to treat my patients better.*”

These qualitative insights illuminate the quantitative pattern of cautious optimism, that is while participants across all career stages recognize AI’s potential to enhance learning, streamline clinical work, and improve patient care, they simultaneously wrestle with profound questions about professional identity, patient safety, accountability, and the preservation of human judgment in an increasingly automated future.

## Discussion

### Key Findings

This study is the first to examine the attitudes and perceptions of medical students and healthcare professionals toward artificial intelligence in Erbil, Iraq. The findings indicate that AI use is widespread among participants, particularly among younger adults and male respondents. Chat GPT is the most commonly used AI among the participants.

Participants were asked to rate their agreement regarding various statements about AI. In general, high scores were noted for statements expressing support for AI development, its necessity for healthcare students and medical professionals, and hope for its role in healthcare going into the future, as well as high scores for statements regarding the need for careful management of AI technology and participants expressing a comparatively low understanding of the ethical implications of AI technology in healthcare. Support and hope regarding the integration of AI into healthcare, as well as thinking that healthcare students should learn the basics of AI were stronger among individuals who used AI.

Participants supported the inclusion of AI education in schools and universities, with especially strong support from those intending to start their own practices or businesses later in their careers. At the same time, concerns about potential job displacement and the risk of physicians becoming overly reliant on AI emerged as the primary reservations.

Qualitative analysis revealed themes of optimism regarding AI and its role in healthcare going forward, with potential improvements in streamlining the work of healthcare workers, saving time to allow healthcare providers to have more time with patients, and improving diagnostics and patient management. However, along with that optimism, a need for caution was expressed by the respondents, who stated concerns regarding patient privacy, handling misinformation, the potential erosion of physician skills in the face of overreliance on AI, the replacement of physicians in healthcare, and potential ethical issues.

### Implications of Findings

The findings have strong implications for both the healthcare system and medical education. The widespread use of AI by both students and professionals alike, especially among the male gender, across all the stages of training, underscores the need to incorporate AI into medical curricula. The high level of support for AI integration in healthcare, together with feelings of hope regarding its potential benefits and understanding its ethical implications, suggests that students are likely to be receptive to formal AI education. Such integration may contribute to greater efficiency in medical education, research, and clinical practice.

In addition, familiarity with AI and prior experience using AI tools were associated with more favorable attitudes toward its adoption. This finding suggests that increased exposure to AI through structured educational programs may help reduce resistance and foster acceptance among individuals who may initially hold negative perceptions of the technology.

The results also indicate that AI may support the development of critical thinking and problem-solving skills, as participants with entrepreneurial ambitions were more likely to endorse the inclusion of AI in medical education. At the same time, concerns regarding job displacement and the ethical implications of AI highlight the need for ongoing conversation about the concerns of healthcare personnel regarding AI during its early stages, ensuring issues such as patient privacy, safety, and personnel replacement are addressed going forward with the incorporation of AI into medical care, as well as the importance of establishing clear regulatory frameworks and incorporating ethical training alongside AI related curricula.

### Literature Comparison

The majority of medical professionals and students in this study (90.8%) reported using AI, and males were more likely to use AI than females. A recent study from Jordan reported the frequency by which medical students used AI for various application, and on average only 22.3% of their participants reported never having used AI for purposes such as exam preparation, assignments, research, brainstorming, etc.^17^ Furthermore, a study from Saudi Arabia showed that 93% of students were familiar with the use of Chat GPT, and that similarly, males used Chat GPT and other forms of AI more than females. ^18^ Furthermore, Duan et al. also reported a gender difference in AI usage, with male students being more enthusiastic than female students.^19^ Hence, the usage rate for AI in Kurdistan is comparable to other parts of the region.

The majority of medical professionals and medical students (85.6%) in this survey think there is a need for AI education in medical curriculums. We found that participants being uninterested in clinical work and those that are looking to pursue their own practice/business were statistically more likely to be in favor of teaching AI in universities. Teng et al reported that 64.8% of their participants thought AI should be part of the medical curriculum.^16^ Furthermore, the responders in their sample that were interested in pursuing research or business had a more positive outlook on AI compared to those that were more interested in pursuing clinical work.^16^ A study conducted in the UK showed that most participants thought AI-related education would be beneficial in their future, and most also agreed medical students need AI-training as part of their curriculum.^20^ A survey conducted in Germany showed that 70.1% of medical students thought AI training needs to be included in their medical education.^21^ Hoffman et al reported that healthcare workers in general lack training regarding AI and see great necessity in AI training, particularly in areas of application and ethical considerations.^22^ Furthermore, comparisons made to regional studies as well, such as Hawal et al, in a study done in Oman on medical undergraduates and faculty, reported a substantial proportion of their students and faculty believing that AI-related training is necessary in medicine, and they also reported opinions that the healthcare system in their country was unprepared for the future integration of AI.^23^ Al-Qerem et al revealed 75% of their participants thought healthcare students should learn the basics of AI.^17^ Thus, healthcare personnel in our region echo international sentiments of feeling unprepared for the future of healthcare regarding AI integration and feeling the necessity of incorporating AI training into medical education.

Regarding participants attitudes towards AI, Teng et al reported an overall positive outlook on the development of AI and its incorporation into healthcare.^16^ The medical students in this sample expressed similar sentiments to the medical personnel and undergraduates in our region with mean scores of around 7 for statements regarding support for AI development and the need for AI education, even higher scores regarding the need for careful management of AI, and a relatively low score for statements of understanding the ethical implications of AI in healthcare.^16^ However, their sample had slightly lower scores for being hopeful about AI and a higher mean scores about believing AI will be impactful in their fields.^16^ This difference could be attributed to the lack of exposure to AI in our region, hence healthcare professionals in our region are less familiar with the potential consequences of AI involvement in medicine and simultaneously may underestimate the impact AI could potentially have on their fields. Physicians in Germany reported mostly positive sentiments towards the involvement of AI in medicine as well, with most believing AI will shape the future of medicine and AI shows high potential to improve patient care, and most participants also thought that AI required thorough scientific evaluation before being implemented in healthcare, and there is a need for training in AI for medical personnel.^24^ Pinto et al reported that most participants agree AI can revolutionize and improve medicine.^21^ Hoffman et al similarly demonstrated that healthcare personnel generally agree that AI can improve patient care, clinical decision making, and healthcare outcomes in their fields.^22^ A substantial proportion of newly graduated Physicians in Saudi Arabia agreed that AI will significantly impact healthcare.^25^ Hence, it seems the internationally positive attitudes towards the future of medicine and AI are reflected by the healthcare professionals and undergraduates of our region.

Regarding concerns for AI, Jackson et al, in a survey in India, examined concerns expressed by medical students and found more than half were worried about patient confidentiality when AI is involved, and a majority expressed that AI could reduce the human aspect of healthcare and adversely affect patient-clinician connections and patient trust.^26^ A Saudia Arabia study on newly graduated physicians revealed apprehension regarding privacy invasions, loss of job opportunities, and worries about the ethical considerations of AI involvement in clinical decision-making.^25^

Regarding the qualitative analysis themes, Teng et al found in their qualitative analysis similar themes of cautious optimism for the future of AI and medicine, as well as themes of worry for being replaced by AI agents^16^. A qualitative survey from the United Kingdom demonstrated that most healthcare professionals thought AI could prove very useful in their fields, and a similar proportion of participants showed concerns about data privacy as well, with a small proportion (10%) of these responders being concerned about their jobs being replaced.^27^ Thus, it seems the attitudes, perceptions and concerns of our locale are a reflection of the international healthcare community at large.

To our knowledge, few studies exist exploring the perspectives and attitudes of healthcare personnel regarding AI and its role in healthcare, particularly in Erbil, hence this data provides valuable insight. In addition, we adopted a multi-centric approach to data collection to provide a more representative sample.

### Limitations

The main limitations of this study are that we collected data only from medical students and professionals based in Erbil, Iraq. This makes the data less generalizable to the entire region of Iraq and its healthcare system as a whole. Furthermore, medical graduates, particularly JHOs, were overrepresented in this survey.

## Conclusion

This survey provides valuable insight into an ever-advancing technology with almost inevitable implications for the future of healthcare. While medical students and physicians alike expressed guarded optimism for the role of AI in healthcare’s future, there was, in general, a dissatisfaction with their understanding of the ethical implications of AI in healthcare. In addition, the need for AI education in healthcare curriculums was highlighted by most participants. These findings highlight the importance of introducing AI into Iraq’s medical education system to better prepare future doctors for its growing role in clinical practice. Potential areas of interest for AI education in medicine could include introductory courses on the technology as a whole, areas of application for AI, and courses on proper security and safety procedure with AI use. In addition, future research could delineate the deficiencies healthcare providers have when it comes to AI to better determine where this education can be best targeted.

## Data Availability

All data produced in the present study are available upon reasonable request to the authors.

## Notes

### Competing Interest Statement

The authors have declared no competing interest.

### Author Declarations

Hawler Medical University, College of Medicine Medical Ethics Commitee granted approval on June 6th, 2025. Meeting code: 9, Paper code: 6

